# Associations between respiratory health outcomes and coal mine fire PM_2.5_ smoke exposure: a cross-sectional study

**DOI:** 10.1101/19002808

**Authors:** Amanda L Johnson, Caroline X Gao, Martine Dennekamp, Grant J Williamson, David Brown, Matthew TC Carroll, Anthony Del Monaco, Jillian F Ikin, Michael J Abramson, Yuming Guo

## Abstract

**Rational:** In 2014, local wildfires ignited a fire in the Morwell open cut coal mine, in south-eastern Australia, which burned for six weeks. Limited research was available regarding the respiratory health effects of coal mine fire-related PM_2.5_ smoke exposure.

**Objective:** This study examined associations between self-reported respiratory outcomes in adults and mine fire-related PM_2.5_ smoke exposure.

**Participants:** Eligible participants were adult residents of Morwell, identified using the Victorian electoral roll.

**Main outcome measures:** Self-reported data were collected as part of the Hazelwood Health Study Adult Survey.

Mine fire-related PM_2.5_ concentrations were retrospectively modelled by the Commonwealth Scientific and Industrial Research Organisation Oceans & Atmosphere Flagship. Personalised mean 24-h and peak 12-h mine fire-related PM_2.5_ exposures were estimated for each participant. Data were analysed by multivariate logistic regression.

**Results:** There was some evidence of a dose-response relationship between respiratory outcomes and mine fire PM_2.5_ concentrations. Chronic cough was associated with an Odds Ratio (OR) of 1.13 (95% Confidence Interval 1.03 to 1.23; p-value 0.007) per 10 μg/m^3^ increment in mean PM_2.5_ and 1.07 (1.02 to 1.12; 0.004) per 100 μg/m^3^ increment in peak PM_2.5_. Current wheeze was associated with peak PM_2.5_, OR=1.06 (1.02 to 1.11; 0.004) and chronic phlegm with mean PM_2.5_ OR=1.10 (1.00 to 1.20; 0.052). Males, participants 18-64 years and those residing in homes constructed from non-brick/concrete materials or homes with tin/metal roofs had higher estimated ORs.

**Conclusions:** These findings contribute to the formation of public health policy responses in the event of future major pollution episodes.

**Key Messages:** *What is the key question?:* Was there an association between mine fire-related PM_2.5_ smoke exposure and self-reported respiratory health outcomes for adult residents of Morwell, approximately 2.5 years after the mine fire?

*What is the bottom line?:* There was some evidence of a dose-response relationship between respiratory outcomes and mine fire-related PM_2.5_ concentrations.

*Why read on?:* There is limited research regarding the health effects of coal mine fire-related PM_2.5_ smoke exposure and to the best of our knowledge, this is the first study to examine self-reported respiratory symptoms associated with smoke exposure from a coal mine fire.

## INTRODUCTION

Coal mine fire smoke contains multiple pollutants known to be harmful to human health, including particulate matter (PM), carbon monoxide, polycyclic aromatic hydrocarbons and benzene,[1, 2]. The chemical profile of pollutants released varies with geographical location, coal composition, meteorology and combustion conditions,[3]. Of these pollutants, fine PM_2.5_, being particulate matter with an aerodynamic diameter <2.5 µm, has been identified as the most harmful to human health,[4]. PM_2.5_ are a mixture of solid and liquid particles created either as primary particles emitted directly into the atmosphere, or secondary particles formed in the atmosphere from pollutant gases.

While multiple studies have found associations between adverse respiratory symptoms and both wildfire,[5-7] and non-wildfire sources of ambient PM_2.5_,[8, 9], limited research is available specifically regarding the respiratory health effects of coal mine fire-related PM_2.5_ smoke exposure,[10]. Mine fire PM_2.5_ has been associated with increased medication dispensing for respiratory health conditions,[11] and studies have found populations living in coal mining communities are at increased risk of respiratory health conditions, even in the absence of fire,[12]. Additionally, as coal mine fires and wildfires are thought to have broadly similar chemical emissions,[10], it is likely the organ systems affected by both would be similar.

In February 2014, local wildfires ignited a fire in the Morwell open-cut coal mine adjacent to the Hazelwood power station and the town of Morwell, in south-eastern Victoria, Australia (Figure 1). The mine fire burned for approximately six weeks. Hourly mine fire-related PM_2.5_ concentrations were estimated to have reached 3700 µg/m^3^, [13] during the initial phase of the fire. The daily average National Environment Protection Measure (NEPM) standard of 25 µg/m^3^ was breached on 27 days during February and March 2014 in the Morwell township,[13]. In response to community concerns following the fire, the Hazelwood Health Study (HHS) was established to investigate the potential long-term health effects of the mine fire on the local population. The current analysis draws upon the Adult Survey component of the HHS, to examine whether there was an association between mine fire-related PM_2.5_ smoke exposure and self-reported respiratory health outcomes for adult residents of Morwell, approximately 2.5 years after the mine fire.

## METHODS

### Study Design & Setting

We conducted a cross-sectional study of self-reported respiratory outcomes following exposure to coal mine fire-related PM_2.5_. The methods have been published elsewhere,[14, 15]. In brief, the study was conducted in the Latrobe Valley, south-eastern Victoria, Australia. The exposure zone comprised five Statistical Area 2 (SA2) districts: Morwell, Yallourn North, Moe, Churchill and Traralgon,[16] (Figure 1). The region was semi-rural and the mine was located on the south-western boundary of the Morwell township. The exposure period was defined as 9 February - 31 March 2014 and the recruitment period was May 2016 - February 2017.

### Participants

Participants were residents of Morwell aged 18 years or older at the time of the mine fire. Eligible participants were identified using the Victorian Electoral Commission (VEC) roll and were invited to take part in the Adult Survey by mail to their last known address. Non-responders were followed up via telephone, mail and public advertisements.

### Variables

Data was collected via a self-report survey, over-the-phone, online or on paper. Variables included participants’ demographic and socioeconomic indicators (age, gender, marital status, education level and employment status). A modified version of the European Community Respiratory Health Survey,[17] was used to identify respiratory symptoms in the previous 12 months and respiratory conditions since 2014.

Participants reported any paid jobs held for at least six months, which may have involved exposure to dust, fumes, smoke, gas vapour or mist. Participants also reported employment in the Latrobe Valley coal mines or power stations, any paid or volunteer positions with the emergency services and, specifically, fire-fighting in the Hazelwood mine fire Controlled Area. Based upon responses, participants were divided into three occupational exposure categories: not exposed; coal mine or power station; exposed but not coal mine power station. Respondents who had ever smoked at least 100 cigarettes or a similar amount of tobacco, in their lifetime were defined as *smokers* as per the World Health Organization (WHO) definition,[18] and categorised as *current* or *former*.

Participants also identified the year of construction of their residence, the main building material, type of roofing and any use of air conditioning during the exposure period. A time-location diary was completed detailing participants’ day and night residential, work and any relocation addresses during the 51 day/night exposure period.

Mine fire-related PM_2.5_ concentrations were retrospectively modelled by the Australian Commonwealth Scientific and Industrial Research Organisation (CSIRO) Oceans & Atmosphere Flagship, using The Air Pollution Model (TAPM v4.0.5), combined with a chemical transport model (CTM),[19, 20]. Modelled data were used due to a paucity of air quality monitoring at the time of the fire,[1, 3, 13]. In particular, no data were available for the first 10 days of the fire in the residential areas of Morwell closest to the mine. The modelling process was described in detail by Emmerson et al.,[13].

Personalised mean 24-h and peak 12-h mine fire-related PM_2.5_ exposure metrics were calculated for each participant based on the addresses listed in their time-location diaries. The Morwell SA2 consisted of 36 SA1s (Figure 2) and exposure was assigned to Morwell addresses at that scale. Exposure was assigned to Churchill, Moe, Yallourn North or Traralgon addresses at the SA2 level (Figure 1). The temporal scale was 12 hours: 6am - 6pm defined as day time exposure and 6pm - 6am defined as night exposure. For each participant, their 51-day and 51-night PM_2.5_ concentrations were averaged to obtain their cumulative mean 24-h exposure metric. Their peak 12-h PM_2.5_ concentration metric was the maximum SA1 or SA2 area level concentration assigned to the addresses recorded in their diary.

### Statistical Methods

Associations between mine fire smoke exposure and respiratory outcomes were investigated for mean and peak PM_2.5_ concentrations using multivariate logistic regression. Analyses were conducted using weighted methods of estimation to reduce the possibility of participation bias. The final multivariate models included exposure variables (mean or peak PM_2.5_ exposure) and common confounders of the respiratory outcomes, namely: age, gender, employment, education, marriage status, smoking, employment exposure, roof materials, building materials, pre-existing asthma and pre-existing chronic obstructive pulmonary disease (COPD). Interactions between PM_2.5_ exposure and both housing and roofing type were tested to rule out the impact of smoke ingress on the true exposure of participants.

Missing data for roofing type and building materials were completed using publicly available real-estate websites and satellite images. Missing observations for remaining variables ranged between 0.06-1.3% (Table S1) and were accounted for using multiple imputation by chained equations (MICE), with 20 iterations,[21, 22].

Overall effects were measured using the odds ratio (OR) for self-reported respiratory outcomes and 95% confidence intervals (CI), associated with a 10 μg/m^3^ increase in mean 24-h and a 100 μg/m^3^ increase in peak 12-h mine fire-related PM_2.5_ smoke concentration. Stratification to assess effect modification by sex (male/female) and age (18-64/65+ years) was also conducted. Statistical analysis was conducted using Stata 15,[23].

### Ethics Approval

The protocol for the Adult Survey was approved by the Monash University Human Research Ethics Committee (Project number 6066). Participants provided informed consent.

## RESULTS

Recruitment results and bias assessment are detailed elsewhere,[14, 15]. In brief, 9,013 Morwell residents were identified by the VEC roll as eligible. Of those, 3,037 (34%) participated, as well as 59 Morwell residents who had not been listed on the roll (N=3096). An assessment of sampling bias found an over-representation of women and older people and differences in smoking patterns. To account for potential bias, the analysis was conducted using weighted methods of estimation (Table 1). Recruitment rates were marginally higher for SA1s closest to the mine (Figure S1).

**Table 1:** Summary of predictor variables across tertile categories of Morwell participants’ mean 24-h mine fire-related PM_2.5_ concentrations (N=3096).

| Predictor Variables | Total | Low Exposure | Medium Exposure | High Exposure |  |
| --- | --- | --- | --- | --- | --- |
|  | Weighted Mean (SD) | Weighted Mean (SD) | Weighted Mean (SD) | Weighted Mean (SD) | p-value* |
| **Age during the mine-fire** | 48.07 (18.59) | 47.71 (18.68) | 46.69 (17.88) | 49.78 (19.05) | 0.020 |
|  | N (weighted %) | N (weighted %) | N (weighted %) | N (weighted %) | P-value* |
| **Male** | 1389 (48%) | 447 (47%) | 461 (50%) | 481 (47%) | 0.371 |
| **Employment** |  |  |  |  | 0.337 |
| Paid employment (FT, PT, self-employed) | 1311 (51%) | 428 (52%) | 441 (53%) | 442 (50%) |  |
| Other (student/volunteer/home-duties/retired) | 1368 (35%) | 464 (36%) | 430 (33%) | 474 (37%) |  |
| Unemployed | 139 (6%) | 50 (6%) | 48 (7%) | 41 (5%) |  |
| Not working due to ill-health | 239 (7%) | 67 (6%) | 82 (7%) | 90 (8%) |  |
| **Highest educational qualification** |  |  |  |  | 0.082 |
| Secondary up to year 10 | 1006 (27%) | 317 (26%) | 357 (29%) | 332 (26%) |  |
| Secondary year 11-12 | 668 (24%) | 203 (22%) | 236 (27%) | 229 (23%) |  |
| Certificate (trade/apprenticeship/technicians) | 996 (34%) | 355 (35%) | 291 (32%) | 350 (35%) |  |
| University or other Tertiary Institute degree | 385 (15%) | 136 (17%) | 113 (12%) | 136 (15%) |  |
| **Married/Defacto** | 1852 (57%) | 676 (61%) | 601 (56%) | 575 (54%) | 0.019 |
| **Smoking status** |  |  |  |  | 0.027 |
| Never | 1495 (51%) | 516 (54%) | 493 (52%) | 486 (47%) |  |
| Former smoker | 1052 (31%) | 356 (30%) | 328 (29%) | 368 (33%) |  |
| Current smoker | 516 (18%) | 141 (15%) | 183 (19%) | 192 (20%) |  |
| **Work exposure** |  |  |  |  | 0.098 |
| Not exposed | 1825 (62%) | 620 (64%) | 611 (64%) | 594 (59%) |  |
| Coal mine/station exposed | 494 (14%) | 162 (13%) | 147 (12%) | 185 (16%) |  |
| Exposed, but not coal mine/station | 777 (24%) | 244 (22%) | 254 (24%) | 279 (25%) |  |
| **Roof main material - iron/tin** | 977 (32%) | 231 (24%) | 346 (34%) | 400 (37%) | <0.001 |
| **Home main material - concrete/brick** | 1927 (61%) | 774 (76%) | 606 (58%) | 547 (51%) | <0.001 |
| **Asthma pre 2014** | 715 (26%) | 248 (27%) | 238 (26%) | 229 (24%) | 0.259 |
| **COPD pre 2014** | 148 (4%) | 44 (3%) | 45 (3%) | 59 (4%) | 0.379 |

\*Weighted chi-square test for proportion differences across exposure categories

Mean 24-h mine fire-related PM_2.5_ concentrations experienced by participants ranged from 0-56 µg/m^3^ with a median of 11 µg/m^3^ and peak 12-h concentrations ranged from 0-879 µg/m^3^ with a median of 132 µg/m^3^ (Figure 3). Peak and mean exposure concentrations had similar distributions whilst peak concentrations had more high value outliers. The Morwell SA1s closest to the mine experienced the highest concentrations, and were also the most highly populated residential areas (Figures 1 and 2).

Potential confounding and outcome variables across tertile categories of mean 24-h PM_2.5_ exposure are shown in Table 1 and Table 2. Exposure for the period 9 February - 31 March 2014 was categorised as: Low: 0-8.6 µg/m^3^, Medium: >8.6-14.1 µg/m^3^ and High: >14.1-56.0 µg/m^3^. Participant age, marital status, smoking status, housing materials and roofing materials varied across exposure categories, which highlighted the need for controlling these variables in the multivariate analysis (Table 1).

**Table 2:** Summary of outcome variables across tertile categories of participants’ mean mine fire-related PM_2.5_ concentrations.

| Outcome Variables | Total | Low Exposure | Medium Exposure | High Exposure |  |
| --- | --- | --- | --- | --- | --- |
|  | N<br>(weighted %) | N<br>(weighted %) | N (weighted %) | N (weighted %) | p-value* |
| Current wheeze | 1317 (42%) | 422 (41%) | 418 (40%) | 477 (46%) | 0.021 |
| Chest tightness | 792 (27%) | 237 (25%) | 265 (27%) | 290 (29%) | 0.201 |
| Nocturnal shortness of breath | 635 (20%) | 203 (20%) | 207 (19%) | 225 (21%) | 0.604 |
| Resting shortness of breath | 611 (20%) | 194 (19%) | 197 (20%) | 220 (22%) | 0.397 |
| Current nasal symptoms | 1358 (44%) | 445 (44%) | 425 (41%) | 488 (47%) | 0.114 |
| Chronic cough | 989 (31%) | 296 (26%) | 340 (32%) | 353 (34%) | 0.004 |
| Chronic phlegm | 785 (25%) | 242 (23%) | 259 (25%) | 284 (27%) | 0.133 |
| Asthma since 2014 | 59 (2%) | 21 (2%) | 18 (2%) | 20 (2%) | 0.722 |
| COPD since 2014 | 60 (1%) | 21 (1%) | 20 (2%) | 19 (1%) | 0.984 |

\*Weighted chi-square test for proportion differences across exposure categories

Multivariate analysis showed a pattern of increasing respiratory symptoms with increasing PM_2.5_, however often CIs were wide and statistical significance was not achieved (Figure 4, Table S2). For each outcome the estimated OR for a 10 μg/m^3^ increment in mean PM_2.5_, was similar to the OR for a 100 μg/m^3^ increment in peak PM_2.5_, and CIs were wider for mean concentrations. The strongest associations were for chronic cough, with ORs of 1.13 (95%CI 1.03 to 1.23; p-value 0.007) per 10 μg/m^3^ of mean PM_2.5_ and 1.07 (1.02 to 1.12; 0.004) per 100 μg/m^3^ of peak PM_2.5_. Current wheeze was associated with peak PM_2.5_, OR=1.06 (1.02 to 1.11; 0.004) and chronic phlegm was associated with mean PM_2.5_, OR=1.10 (1.00 to 1.20; 0.052). Those with pre-existing asthma or COPD reported worse respiratory symptoms than those without (Table 1), but no interaction with exposure was identified. A comparison using MICE and complete case analysis yielded essentially the same results.

The sex stratified analyses suggested that estimated ORs were generally higher for males compared with females (Figure 5, Tables S3-S4). The highest ORs were observed in men with asthma since 2014, OR 1.58 (1.10 to 2.29; 0.014) for mean PM_2.5_ and 1.43 (1.14 to 1.78; 0.002) for peak PM_2.5_, with little evidence in women. Among men, the ORs for chronic cough were estimated as 1.20 (1.05 to 1.37; 0.007) for mean PM_2.5_ and 1.07 (1.00-1.14; 0.050) for peak PM_2.5_ and current wheeze 1.10 (1.03 to 1.17; 0.004) for peak PM_2.5_. Amongst females, the highest ORs were for chronic cough, OR 1.06 (1.00 to 1.12; 0.051) for peak PM_2.5_. Age stratification showed higher estimated ORs for participants aged 18-64 years, compared with those 65 and over (Figure 6, Tables S5-S6). For those 18-64, the highest ORs were observed for chronic cough, OR 1.17 (1.04 to 1.30; 0.008), and chronic phlegm, OR 1.14 (1.02 to 1.28; 0.023), both for mean PM_2.5_. For those 65 and over all CIs incorporated the value one.

A protective effect of brick/concrete housing materials was identified in the multivariate regression models for both mean and peak PM_2.5_, for wheeze, cough and nocturnal and resting shortness of breath symptoms (Table S7). Two-way interaction analyses of home building materials and PM_2.5_ concentrations, found associations between nocturnal and resting shortness of breath outcomes and the interaction variable for mean PM_2.5_ (Table S8). Estimated ORs between PM_2.5_ and respiratory symptoms were higher for participants with non-brick/concrete housing compared with brick/concrete housing (Figure S2). For participants with metal/tin roofing materials, the multivariate regression models estimated increased odds for both mean and peak PM_2.5_, for nasal symptoms and phlegm (Table S7). No interaction was identified between roofing materials and PM_2.5_ concentrations.

## DISCUSSION

To the best of our knowledge, this is the first study to examine self-reported respiratory symptoms associated with smoke exposure from a coal mine fire. Our findings showed some evidence of a dose-response relationship between mine fire-related PM_2.5_ smoke concentrations and self-reported respiratory outcomes collected about 2.5 years after the fire. The strongest relationship observed was between chronic cough and mean PM_2.5_ exposure. Chronic cough and current wheeze were associated with peak exposure, and chronic phlegm with mean exposure. Males, participants aged 18-64 years and those residing in homes constructed from non-brick/concrete materials or homes with tin/metal roofs had higher estimated ORs.

Our analysis builds on the HHS Adult Survey volume 2 report,[14], which found an association between chronic cough and tertile categories of participants’ mean 24-h mine fire-related PM_2.5_. In our analysis, building and roof material types were included as additional predictor variables, age and gender stratification were undertaken, peak 12-h exposure was examined and exposure was analysed as a continuous variable. It is possible these factors have improved the statistical power of the model, facilitating the identification of additional associations between respiratory outcomes and mine fire-related PM_2.5_ smoke exposure.

Direct comparisons between our study and existing peer-reviewed literature are difficult, given the limited published research regarding coal mine fire smoke exposures. Previous research conducted by the HHS showed an association between the dispensing of respiratory medications in the Latrobe Valley and mine fire-related PM_2.5_ smoke exposure,[11] and some comparison is possible with other studies that have investigated wildfire PM smoke exposures. Associations were found between wildfire PM_10_ exposure following an American wildfire and survey-reported respiratory symptoms in children,[24]. An Australian study found an increased risk of emergency department attendance for asthma during a 2006/7 wildfire,[6]. Several Canadian studies have found associations between wildfire PM_2.5_ and asthma-related physician visits,[25-27], hospital admissions,[25] and the dispensing of the reliever medication salbutamol,[26-28].

Cough is a physiological mechanism to clear inhaled particles from the respiratory tract and phlegm an indicator of mucus production, so the associations found between PM_2.5_ exposure and these symptoms are plausible. The finding that wheeze was more strongly associated with peak exposure compared to mean exposure, may be because residents more prone to respiratory conditions were more likely to take protective action such as relocating away from the fire,[24] or increasing inhaled medications. Mine fire-related PM_2.5_ concentrations peaked on day two of the fire,[13] at which point residents may still have been residing at their Morwell residential or work address and their peak exposure value would reflect this. If participants subsequently relocated, they would have a reduced mean exposure score making it more difficult to identify an association.

Stronger dose response relationships were observed in males and those aged 18-65 years. Possibly these groups were more active outdoors and/or employed during the mine fire and had less opportunity to relocate outside the exposure zone. Additionally, there may be an element of survivor bias,[29] in the older age group. Relocation may also explain why gender stratification found chronic cough in females was more strongly associated with peak compared to mean exposure, as women may have had more flexibility to relocate. The stronger OR between PM_2.5_ and asthma since 2014 in males may reflect a combination of the mine fire triggering the diagnoses of previously unrecognised asthma and/or small number instability (n=18) as reflected in the wider CIs [OR 1.58 (1.10 to 2.29; 0.014)].

There is mixed evidence in the literature regarding the age group most susceptible to PM related respiratory symptoms. Stronger associations were found in the 20-34 years group, relative to other ages, for the dispensing of respiratory medications following the Hazelwood mine fire,[11]. Henderson et al.,[25] found stronger associations for middle aged adults relative to older adults for respiratory physician visits following a wildfire. However, other wildfire studies found the relative strength of associations for different age groups varied for different respiratory outcomes,[7, 30]. Gender stratification has generally found women have stronger associations than males. Following the mine fire, we found women had a slightly stronger association than men for the dispensing of respiratory medication,[11]. Following wildfires, an increased risk of hospital admissions for asthma in females was reported by Delfino et al.,[7] and mixed results for different respiratory-related emergency department visits were found by Tinling et al.,[31].

Building materials of participants’ residences were associated with respiratory outcomes. Residences constructed from brick/concrete were protective, possibly because relative to weatherboard houses there was reduced penetration of PM_2.5_. Residences with roofs constructed from tin/metal were associated with increased ORs, possibly because in the study area houses with tin roofs were often constructed from weatherboard and houses with tile roofs from brick. While metal roofs may have reduced PM_2.5_ penetration rates relative to tiles, the weatherboard house construction would have increased penetration.

Wildfire studies have found particulate penetration rates are higher for homes with reduced air-tightness,[32, 33] and that reducing indoor PM_2.5_ concentrations with air-filters reduces respiratory symptoms,[34]. There may also have been some inter-relationship with socio-economic factors. Weatherboard houses were generally less expensive and not always well maintained and studies have found lower socio-economic groups may be more susceptible to wildfire smoke exposure,[35, 36]. The two-way interaction found between PM_2.5_ and housing materials for shortness of breath may be a combination of socio-economic factors, pre-existing health conditions and possible indoor exposure differences. Nocturnal shortness of breath can be an indicator of asthma or heart failure and resting shortness of breath an indicator of severe COPD, and those from lower socio-economic groups may be more prone to illness. While our study controlled for known socio-economic and health confounders, it is possible some residual confounding remained. These study results contribute to health policy responses in the event of future mine fire or wildfire pollution episodes. Study participants included both males and females and adults of all ages, and therefore the results would have external validity in other similar communities affected by pollution episodes.

### Strengths and Limitations

A major strength of our study was the ability to gather individual level health data, confounding factors, including residence construction materials, detailed hourly mine fire-related PM_2.5_ concentrations and time-location dairies for each participant. Additionally, the use of individual exposure scores, rather than community level exposure measurements may have reduced the risk of exposure misclassification and increased the power of the study to identify associations between health and exposure,[37]. However, some uncertainties were associated with the use of modelled PM_2.5_ concentrations, such as assumptions regarding emission characteristics, coal burn depth, local wind conditions and smoke plume dispersion,[13]. Further, details regarding housing material and roofing type were only collected for the principal residence of each participant and the proportion of time spent at this location would have varied between participants.

There was a 26-month lag between the end of the mine fire (March 2014) and the commencement of recruitment (May 2016) which may have led to some recall bias. Most respiratory symptom questions referred to the previous 12 months, which would at minimum have been 14 months after the fire period. Therefore, influences other than the mine fire may have contributed to symptom reporting. Alternatively, it is possible the reporting of respiratory symptoms after an extended period post the mine fire, may be an indicator of persistent or long-term impacts of the event. Respiratory outcomes may also have been overestimated due to comorbidity between respiratory symptoms and stress. Traumatic stress has been associated with the activation of a number of neurobiological systems, including inflammatory cytokines,[38], which may in turn impact on airways reactivity,[39, 40] and therefore stress may have presented as respiratory outcomes.

## CONCLUSION

This study found dose response relationships between increments in coal mine fire-related PM_2.5_ smoke exposure and increases in respiratory outcomes in adults, including cough, phlegm and wheeze, about 2.5 years after the event. The strongest associations were observed for males, for adults under 65 years and participants living in non-brick/concrete residences. These findings contribute to the formation of public health policy responses in the event of future major pollution episodes.

## Data Availability

Deidentified data could be made available following an individual approach to the authors, approval by the Project Steering Committee and an institutional ethics committee.

## ACKNOWLEDGEMENTS

The Hazelwood Health Study is a large program of work that comprises a number of research streams in addition to this Adult Survey stream. Those research streams are run by a multidisciplinary group of academic and professional staff from several Institutions including Monash University, the University of Tasmania, Federation University, University of Adelaide and the Commonwealth Scientific and Industrial Research Organisation. All of these staff are thanked for their contribution to this collaborative work. We also thank all HHS participants.

---

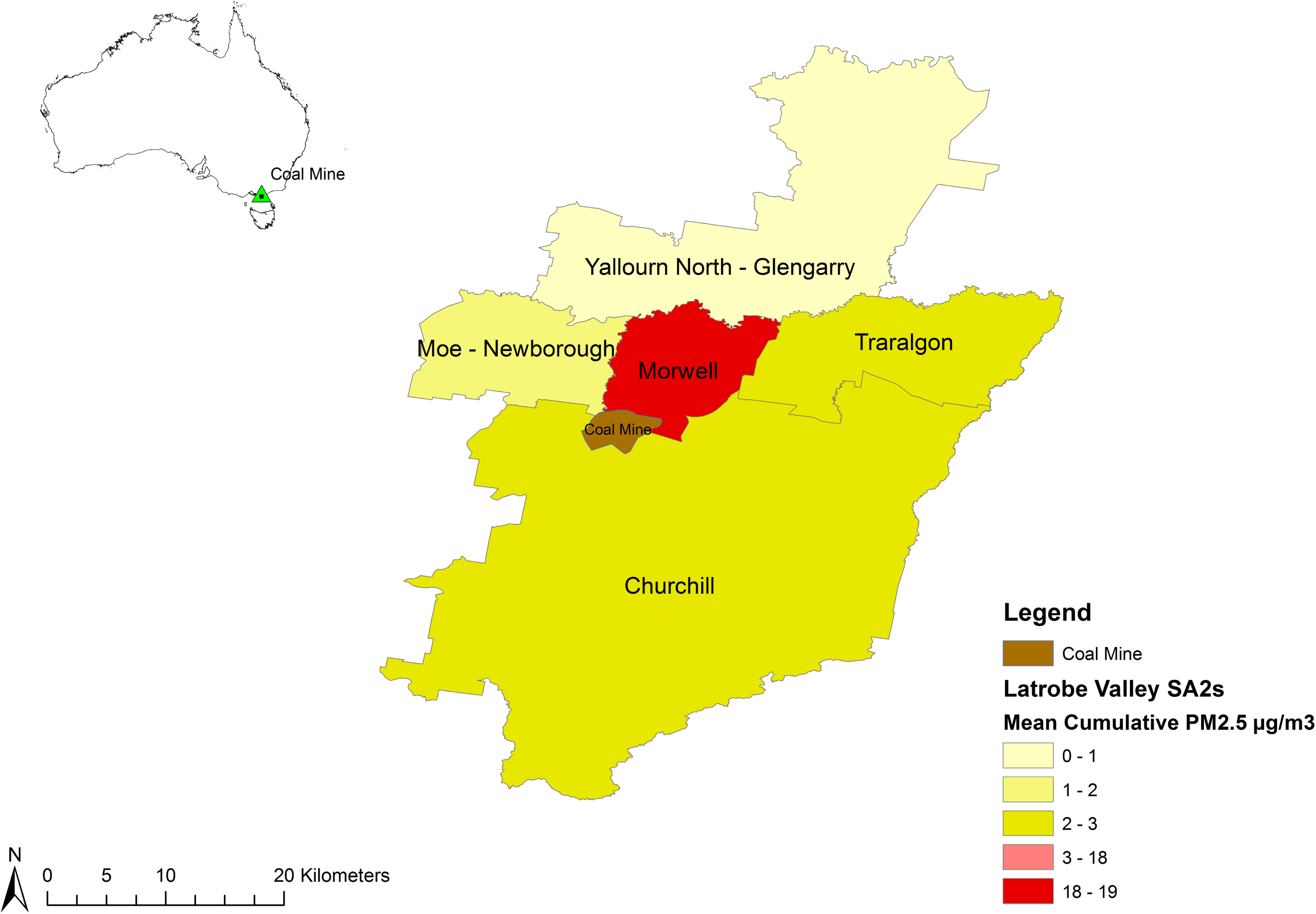

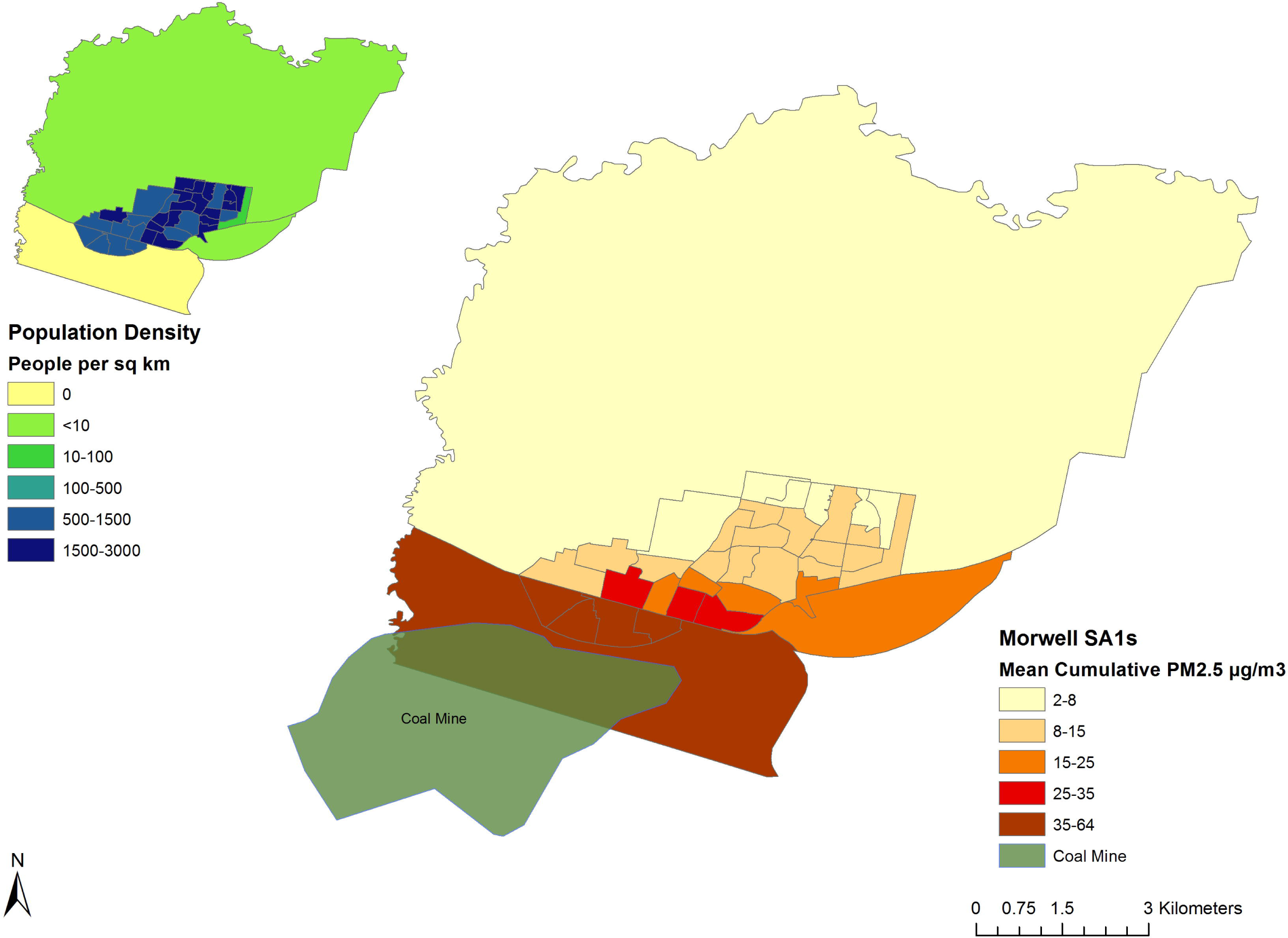

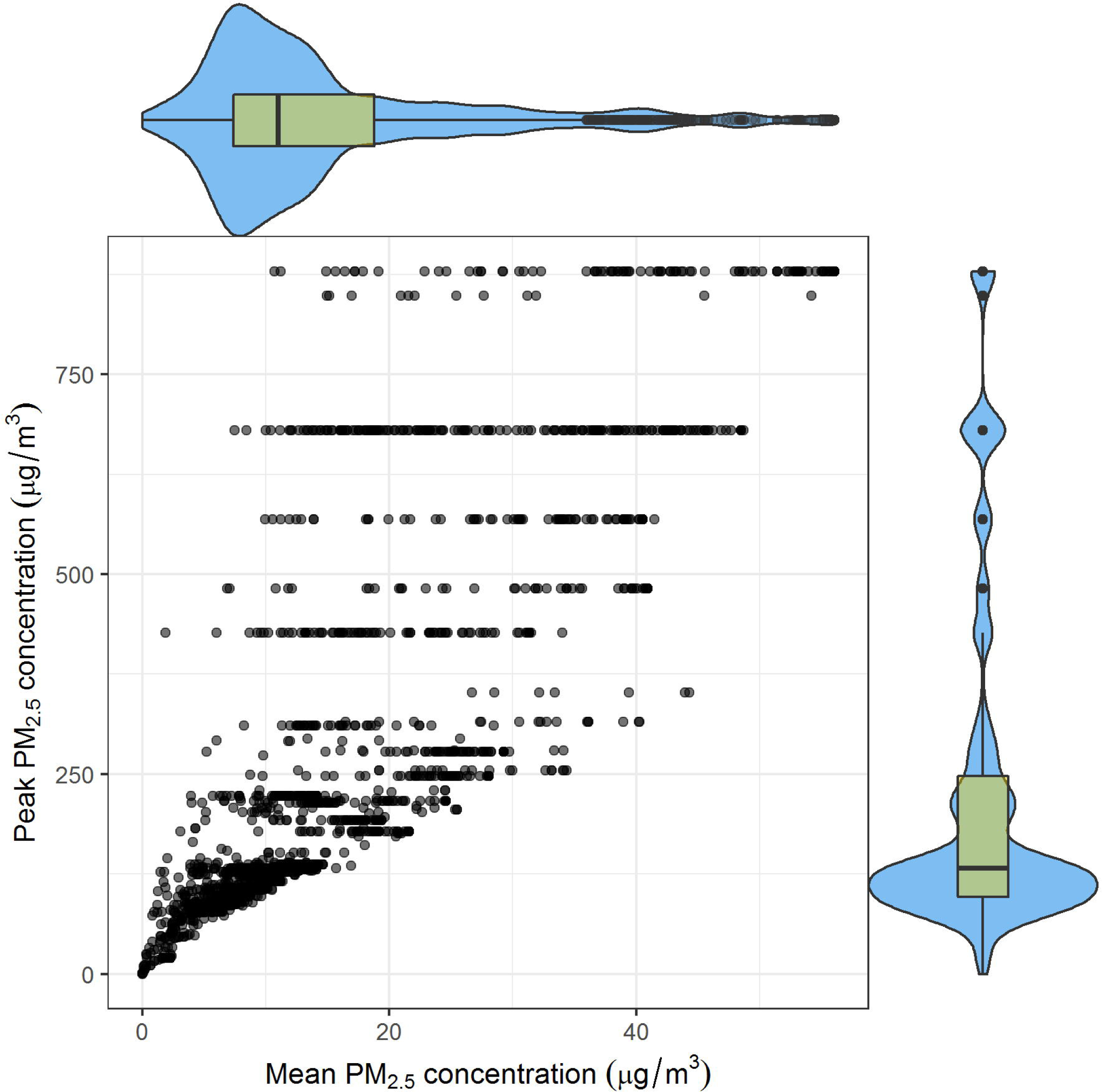

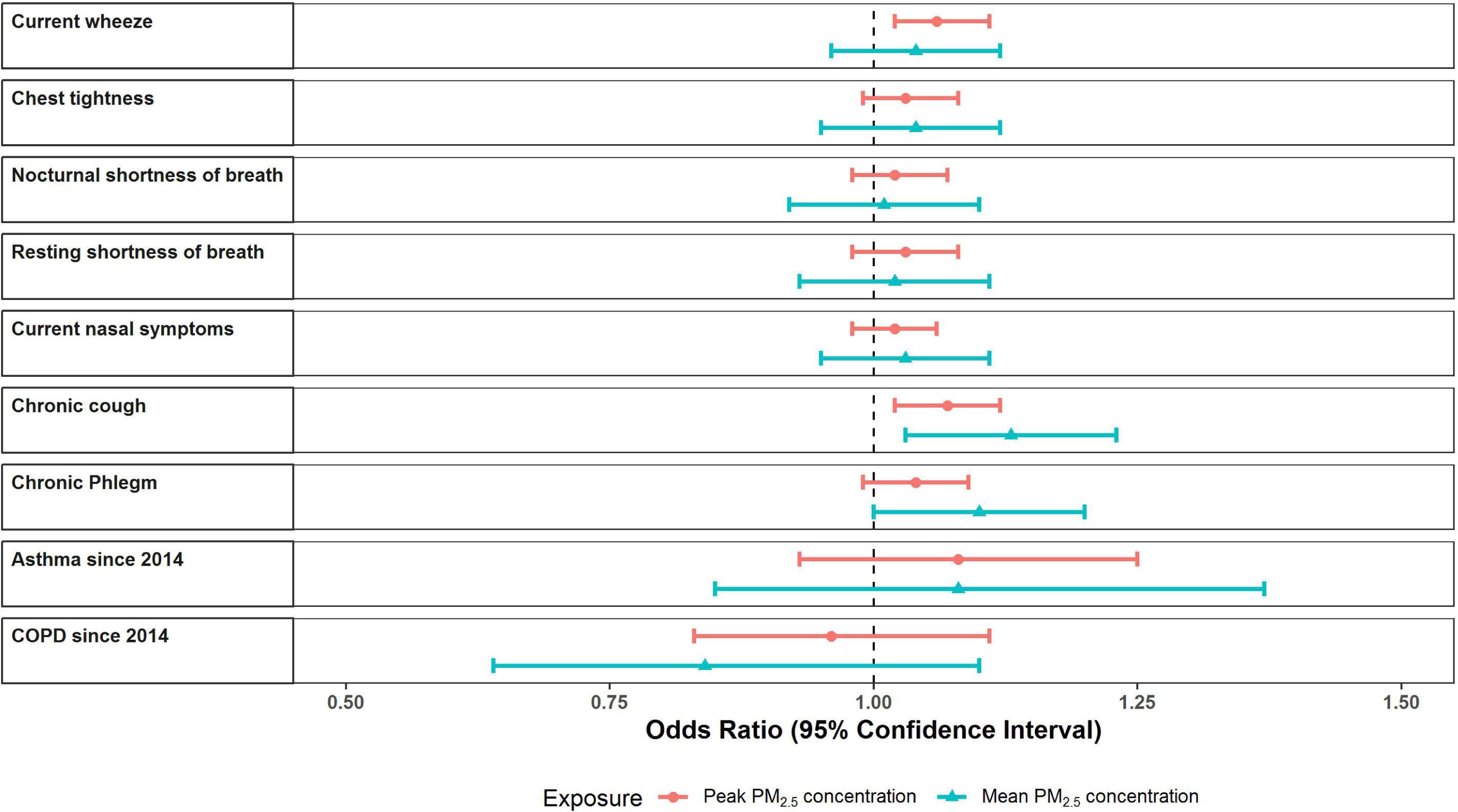

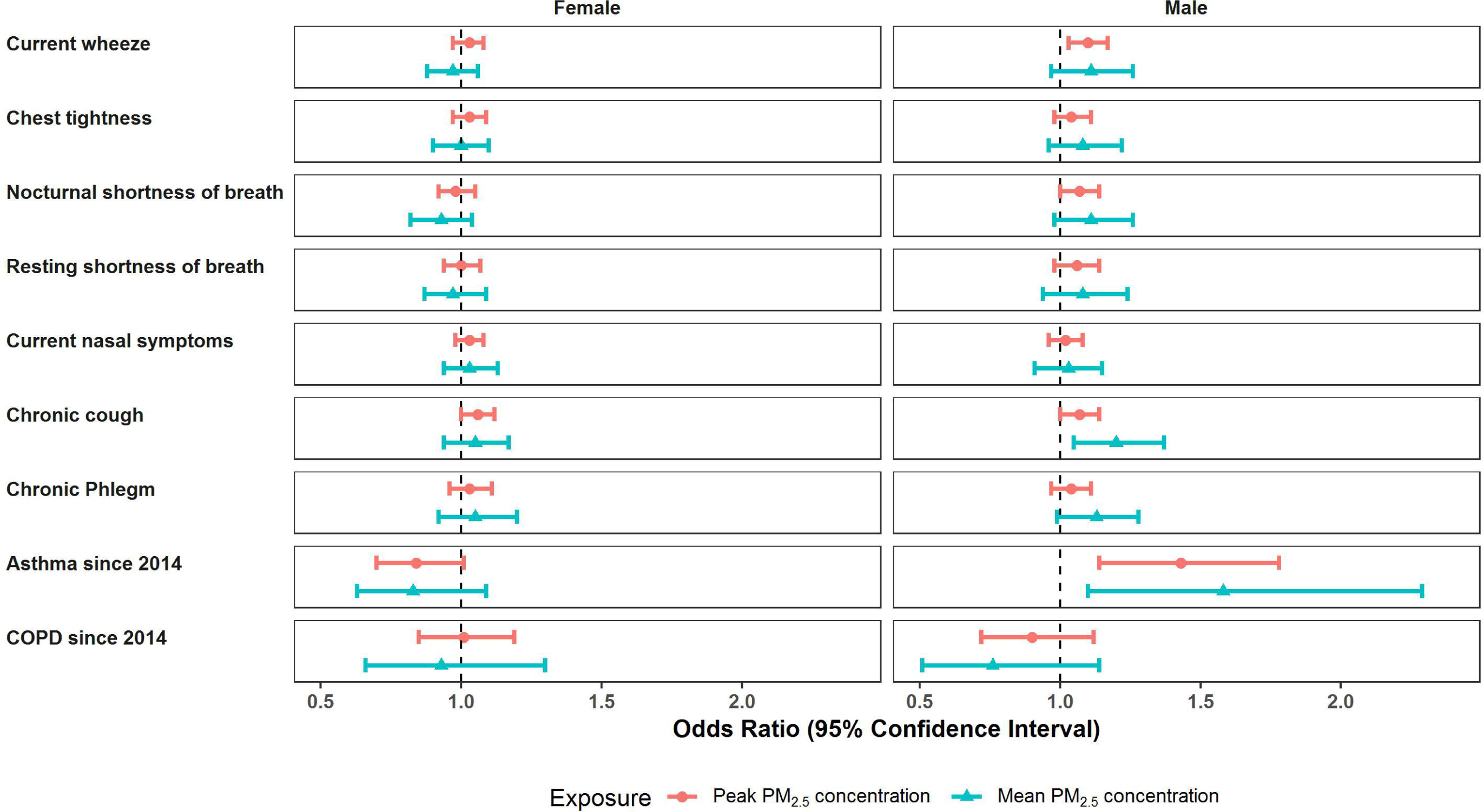

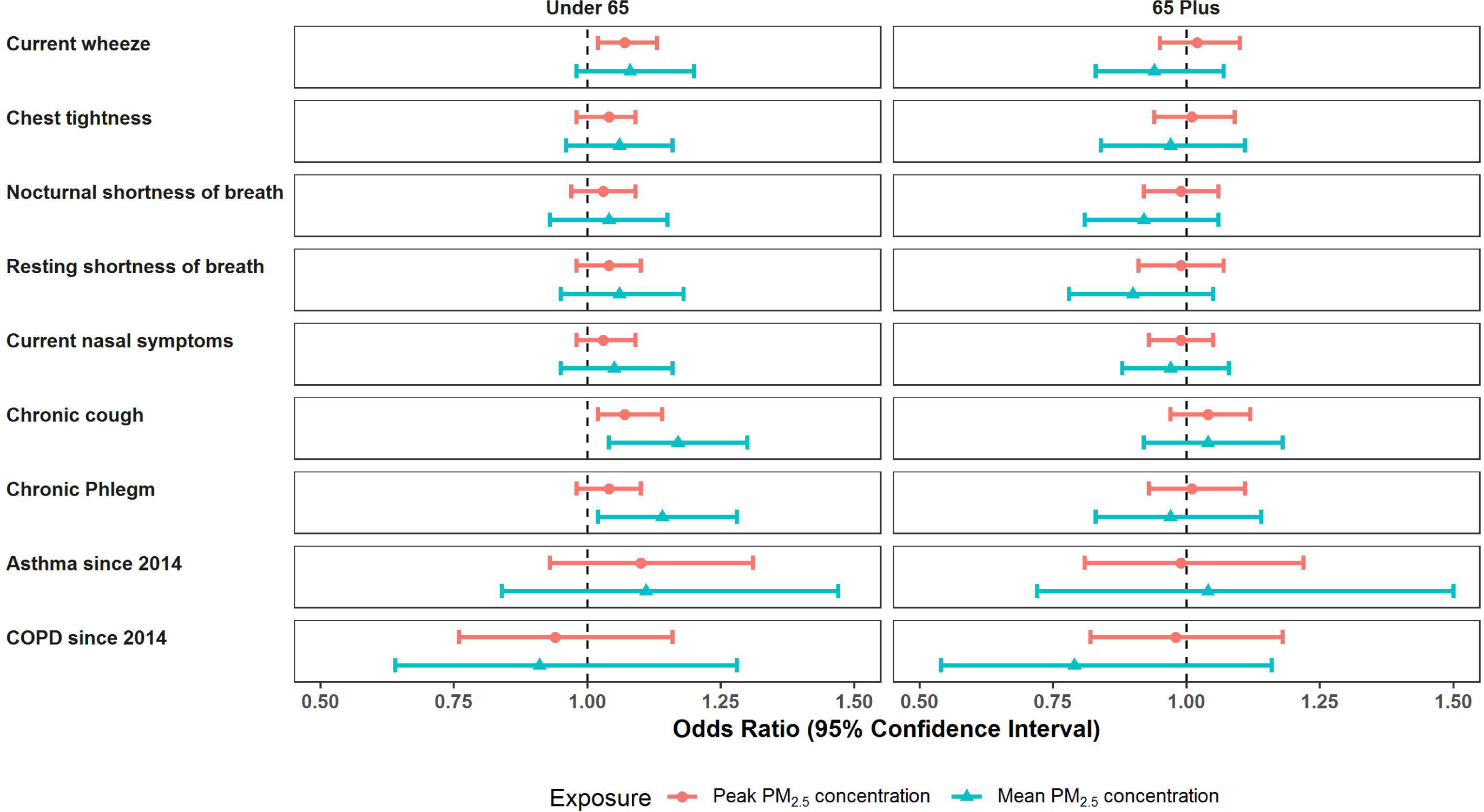

